# Understanding the causes and consequences of low statin adherence: evidence from UK Biobank primary care data

**DOI:** 10.1101/2025.01.23.25321011

**Authors:** Deniz Türkmen, Xiaoran Liang, Jane A H Masoli, Dipender Gill, Luke C Pilling, Jack Bowden

## Abstract

**Background:** Statins are commonly prescribed to lower LDL cholesterol. Clinical guidelines recommend 30-50% reduction within 3 months, yet many patients do not achieve this. We investigated the impact of patient characteristics and genetics on LDL-c reduction, treatment adherence, and adverse clinical outcomes.

**Methods:** We analysed 76,000 UK Biobank participants prescribed atorvastatin or simvastatin in primary care: 41,000 had LDL-c measurements before statin initiation (median=16 days prior, IQR=28) and within a year of starting treatment (median=89 days, IQR=125). Adherence was defined as the “proportion of days covered” (PDC). We estimated associations between PDC within one year of statin initiation, genetic factors, post-treatment LDL reduction, and clinical adverse outcomes. For 13,000 patients with ≥3 LDL-c measures, we used inverse probability weighting methods to estimate the effect of sustained adherence intervention on LDL-c reduction longitudinally.

**Results:** Predictors of LDL-c reduction following statin initiation included the time until the 1^st^ measurement, PDC, and the pharmacogenetic variant *SLCO1B1*\*5. LDL-c reduction was greater in those with high adherence versus lower adherence (38% reduction when PDC>95% [high] vs. 15% when PDC<50% [low]). Longitudinal causal modelling showed that the most recent PDC measure exerted the largest influence on overall LDL-c reduction, followed by the initial PDC.

Genetic predictors of reduced PDC included liability to schizophrenia (Coef_top 20%_-1.94, 95%CI -2.69 to -1.19), whilst genetic liability to cardiovascular diseases increased PDC (Coef_top 20%_1.30, 95%CI 0.55 to 2.05). High PDC was associated with increased risk of incident iron deficiency anaemia (HR 1.30, 95%CI 1.09-1.54) and cataract (HR 1.20, 95%CI 1.07-1.34), and decreased risk of incident coronary heart disease (HR 0.78, 95%CI 0.73-0.84).

**Conclusion:** We identify substantial variability in the time to first on-treatment LDL measurements and also in adherence to statin medication, highlighting a gap between NHS guidelines, LDL monitoring and statin adherence. We show its subsequent impact on long term health, demonstrating the potential effect of targeted interventions to improve adherence. We identify important predictors of reduced statin effectiveness, including pharmacogenetic variants, polygenic scores, but most of all, adherence. Tailored statin therapy strategies with patient education on statin indication and adherence could optimise treatment efficacy, safety, and long-term clinical outcomes.

## Introduction

Cardiovascular diseases (CVDs) remain the leading cause of mortality and morbidity in adults globally (1). Low-density lipoprotein cholesterol (LDL-c), a key modifiable risk factor, can be lowered with statins, reducing CVD risk by ∼22% per 1 mmol/L (18mg/dL) reduction (2). National guidelines, including the UK National Institute for Health and Care Excellence (NICE) and the American Heart Association (AHA), recommend a 30-50% target LDL-c reduction for those aged 40-75 with a high CVD risk with a follow-up lipid check at 2-3 months post-statin initiation (3–5). In the UK, for secondary prevention, the target LDL-C is <2 mmol/L (36mg/dL). Many individuals with detected hypercholesterolaemia are not followed up in the desired time frame and do not achieve these targets (6–9). Adherence to statin therapy in real-world studies can be <50% within the first year of treatment (10–12), far lower than in randomized controlled trials (RCT) conducted with close clinical monitoring and mostly concentrating on patients after an CVD event or hospitalization (13,14). Poor long-term adherence to statin therapy is associated with higher hospitalization rates (15), yet the patient-related causes and consequences of statin adherence in routine primary care remains unclear.

Recent trials (16,17) show those at high genetic risk for CVD experience greater benefits from statins in primary prevention, but whether this effect is pharmacogenetic or simply an artefact of increased pre-treatment LDL levels is unclear. Understanding any effect of pharmacogenetic variants and polygenic scores (aggregated over multiple genetic loci) on statin adherence and LDL-c reduction is of vital importance, since they offer the potential for stratified medicine to better support the identification and optimization of treatment outcomes in high-risk populations. Recent largescale studies have investigated socio-demographic and genetic risk factors of statin adherence (18), or genome-wide association studies of lipid response to statins (19), but have not considered adherence, LDL-c reduction, and adverse clinical outcomes in parallel. To address this, we conducted an extensive analysis of 76,000 UK Biobank participants prescribed statin medication in the linked primary care records. We estimated the impact of time to first on-treatment follow-up, pharmacogenetic variants, polygenic scores, and adherence on LDL-c reduction within 1 year of initiating statin treatment. We also estimated the impact of low adherence on adverse clinical outcomes. To further evaluate the effect of interventions to increase statin adherence on LDL levels over time, we performed longitudinal causal modelling of individual patient LDL trajectories, paired with inverse probability weighting (IPW) methods (20) to control for observable time-varying confounders of adherence and LDL.

## Methods

### Cohort: UK Biobank

The UK Biobank (UKB) recruited 503,325 community-based volunteers aged 40-70 from Wales, Scotland, or England over the period 2006-2010. Participants provided blood samples for genetic and biochemical analyses. This study involves two analyses: 1) using the linked primary care data from General Practice (GP) available in 230,096 participants (45.7% of UKB) (Figure 1) to examine LDL reduction, and 2) Incorporating secondary care data (hospital) with the GP data for the incident outcomes.

**Figure 1.**
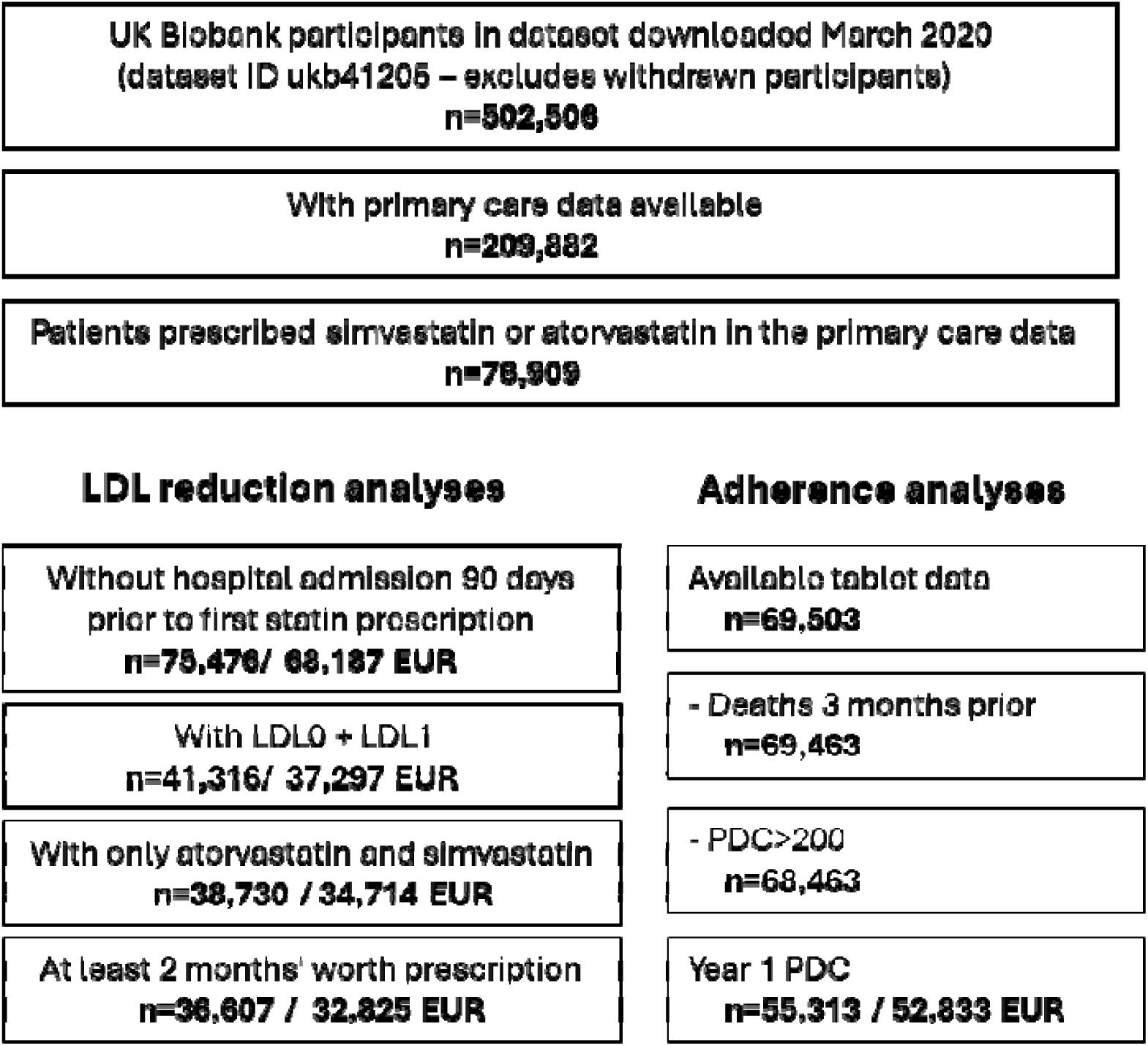
Flowchart of the UK Biobank cohort. LDL0: pre-statin LDL-c measurement, LDL1: post-statin LDL-c measurement, EUR: Genetically European ancestry

GP data includes drug name, date of prescription, number of tablets, and drug code (in Clinical Read version 2, British National Formulary (BNF) or Dictionary of Medicines and Devices (DM+D) format, depending on provider) and are up to September 2017 (EMIS/Vision system in Wales) and August 2016 (TPP system supplier in England). We included simvastatin and atorvastatin prescriptions, classifying first doses by NICE guidelines (21) : 8% low-intensity, 76% medium-intensity, and 16% high-intensity in the UKB cohort (Appendix Table 1 and 2).

**Table 1.**
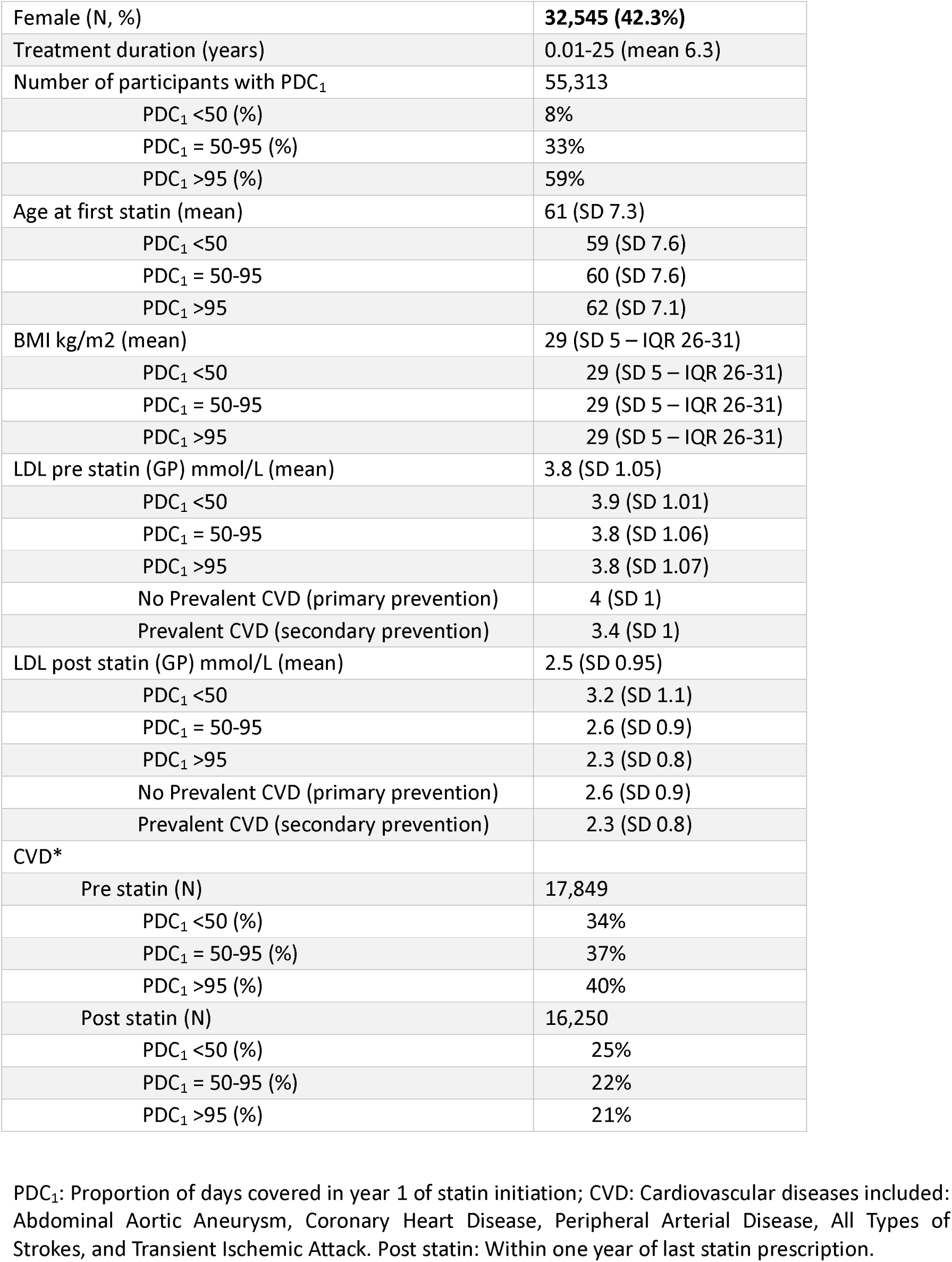
Descriptive table of patients who have GP prescribed statin.

We identified primary and secondary care-diagnosed CVDs (Abdominal Aortic Aneurysm, Coronary Heart Disease, Ischemic Stroke, Ischemic Stroke or Transient Ischemic Attack, Peripheral Arterial Disease, All Types of Stroke, and Transient Ischemic Attack) based on clinical guidelines for initiating statin treatment for secondary prevention (22). Supplementary Table 1 for the ICD-10 codes.

### Genetic Data

UKB obtained 805,426 directly genotyped variants via the Affymetrix Axiom UKB array (in 4381427 participants) and the Affymetrix UKBiLEVE array (in 49,950 participants). Imputation was performed by the central UKB team for 487,442 participants and ∼96 million genetic variants were obtained. Neither participants’ nor their healthcare providers received genotype data as part of the study, so this information could not directly influence treatment. 451,367 (93%) participants genetically similar to the 1000Genomes EUR reference population (“European-like”) were included in our genetic analyses to minimise the effect of population stratification (this was identified via Principal Components Analysis using the 1000Genomes reference panels, see (23) for details).

We also included polygenic scores (PGS) derived by Genomics PLC (24) (UKB category 301): 8 PGS for relevant biomarkers and patient characteristics (e.g., LDL, BMI) and 28 PGS for genetic liability to disease (e.g., CVD, schizophrenia). See Supplementary Table 2 for the details.

### Adherence as measured by Proportion of Days Covered (PDC)

For each patient with a history of statin treatment initiation as well as pre- and post-treatment LDL measures (n=69,503), we sought all available prescribing data. We then calculated the number of tablets prescribed to each individual in a given time period, to create the variable:

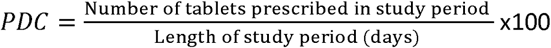

We first focused on adherence in the first year of treatment, which we call *PDC_1_*. If patients received a new prescription that extended past the first 12 months, we included the time until the next prescription in the ’length’ calculation for *PDC_1_*. However, if there were no subsequent prescriptions, we excluded that prescription from the calculation (n=7,446). We also excluded patients who died within 3 months of starting statins (n=39) and those with a *PDC_1_* >200% (n=714). See Figure 1 for a flowchart of this process and the final number of included participants.

We used a three-category definition for *PDC_1_* when treating it as the exposure: <50% , 50-95% , >95% based on observed adherence patterns in the cohort and partly in line with previous research [e.g., for <50%] (12). In addition, when treating it as the outcome variable and estimating its predictors via multivariable regression modelling, we used the original (continuous) *PDC_1_* measure. When performing longitudinal causal modelling of individuals’ LDL trajectories over three post-treatment follow up visits, continuous measures for PDC adherence over participant-specific study windows were used as covariates and in the derivation of inverse probability weights.

### Outcomes

#### LDL change in 1 year

We obtained GP-recorded LDL measurements for patients on statins (see Supplementary Table 3 for the read codes used). We identified the baseline LDL measure (within 180 days and preceding first prescription) and the first follow-up measure following statin initiation, restricting analyses to participants with a follow-up measure within 12 months of statin initiation. LDL measurements for patients prescribed their first statin within 90 days of hospital discharge for a cardiovascular event were excluded, as these prescriptions may not have been recorded by the GP. Extreme measurements (LDL <1 mmol/L: n_LDL0_=1,402, n_LDL1_=2,081 and LDL >8 mmol/L: n_LDL0_=29, n_LDL1_=13) were excluded (see Figure 1). We defined an LDL change measure as the post-statin/pre-statin LDL ratio:

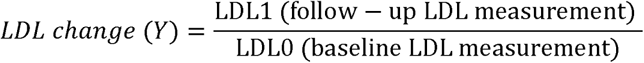

#### Long-term clinical outcomes

Eighty four long-term clinical outcomes were investigated to identify intended and unintended adverse events. This encompassed all common chronic diseases lasting more than 3 months with a prevalence of >0.5% in older adults (25).

### Analyses

Figure 2 provides a detailed visualisation of our analysis strategy. All models in Analysis 1 are adjusted for sex, age at first statin prescription, the baseline assessment centre, first dose and education unless stated otherwise.

**Figure 2.**
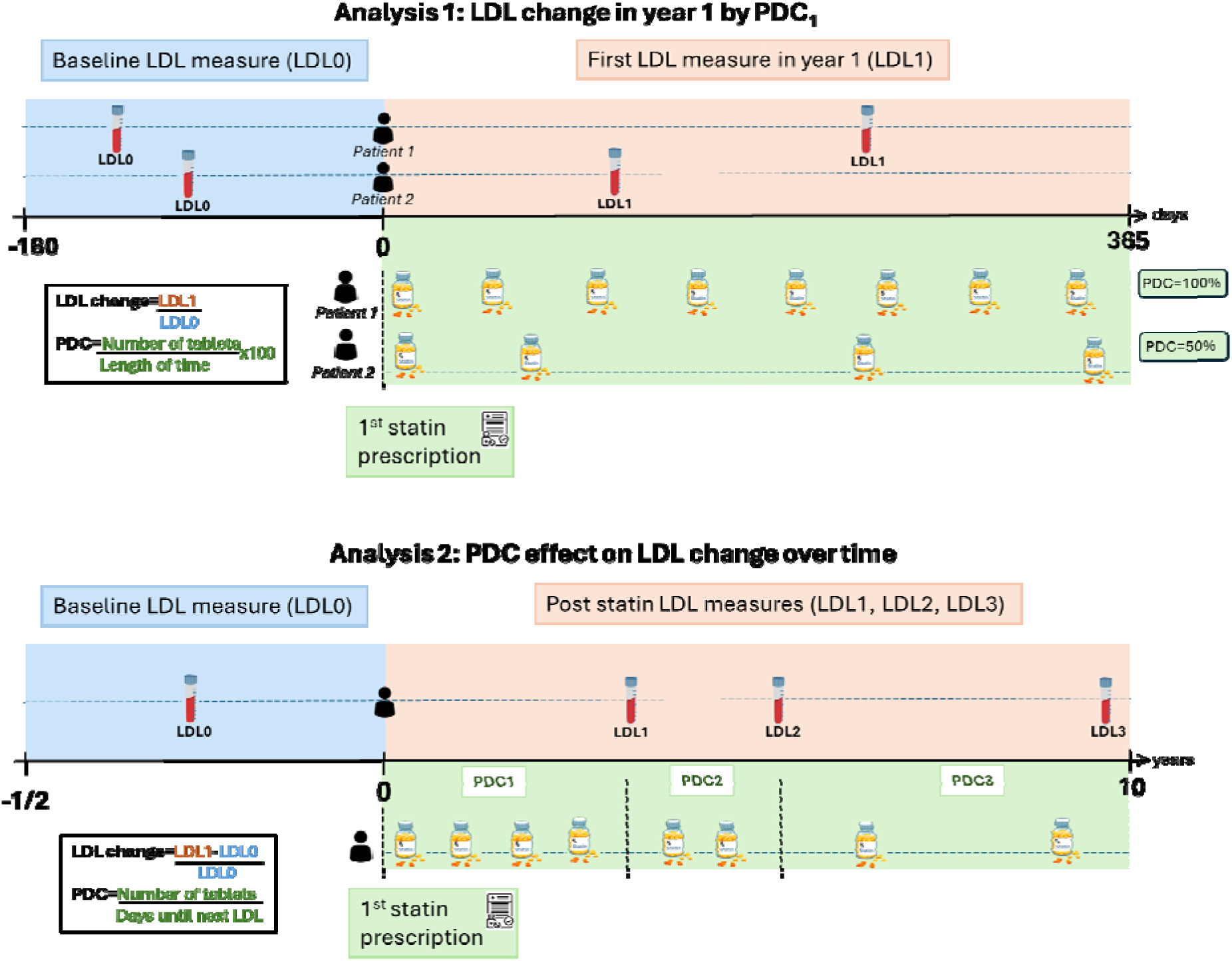
Design of the analyses. For each patient with a history of statin treatment initiation as well as pre and post treatment LDL measures (n=69,503), we sought all available data on the date and quantity their prescription. We then calculated the number of tablets prescribed to each individual in a given time period, to create the PDC variable. <u>Analysis 1</u>: We estimated LDL change for varying the adherence in the first year of treatment, which we call. If patients received a new prescription just before the end of year 1, we included the time until the next prescription in the ’length’ calculation for. However, if there were no subsequent prescriptions, we excluded that latest prescription from the calculation (n=7,446). We also excluded patients who died within 3 months of starting statins due to pre-existing illness (n=39) and those with a >200% (n=714). <u>Analysis 2</u>: We attempted to evaluate how intervening to increase statin adherence would affect a person’s LDL change over a longer sustained period, adjusting for fixed confounders of PDC and LDL, as well as time-varying confounding of previous PDC and LDL measures on their future values. For patients prescribed statins, we included individuals with at least three post-statin LDL measures over a ten-year window for the analysis (n=13 315).

### Analysis 1: Cross-sectional analysis of LDL change

#### LDL change explained by week of follow-up

We tested associations between LDL change and the time interval between their pre- and post- treatment measurements, using a linear regression model, adjusted additionally for dose of first atorvastatin and simvastatin prescribed in year 1 and the prevalent cardiovascular disease diagnoses. Doses were organised into a categorical variable based on NICE guidelines (21) (Appendix - Table 1). We repeated this analysis in the <=95% and >95% *PDC_1_* groups separately.

#### Modelling LDL change by genotype, PDC and time of follow-up measure

We next tested associations between LDL change and genetic variant rs4149056 using a linear model that allowed for a quadratic effect of time-to-follow-up on treatment in European-like participants and an interaction between time on treatment and rs4149056. We also conducted a linear regression analysis to test the associations between polygenic scores and LDL change. We additionally adjusted for 10 genetic principal components (PCs) in these two analyses.

Next, we tested the association between LDL change and PDC_1_ using the same model, but with PDC in place of rs4149056. This analysis did not adjust for genetic PCs but accounted for the prevalence of CVD before statin use, including abdominal aortic aneurysm, atrial fibrillation, coronary heart disease, peripheral arterial disease, all types of stroke, and thromboembolic disease. For further technical details on the statistical modelling see the Appendix.

### Analysis 2: Longitudinal analysis on LDL trajectories

#### Causal modelling of hypothetical LDL change via sustained PDC intervention

We applied and refined causal modelling approaches to evaluate how intervening to increase statin adherence would affect a person’s LDL reduction over a longer sustained period, adjusting for fixed confounders of PDC and LDL, as well as time-varying confounding of previous PDC and LDL measures on their future values. For patients prescribed statins, we included individuals with at least three post-statin LDL measures over a ten-year window for the analysis (n=13,315). We defined LDL reduction (LR) as the percentage change between each of their post-statin LDL measures and the pre-statin baseline level (denoted by LDL_O_) as

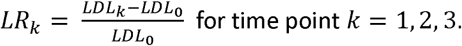

Figure 3 shows the distribution of patients’ three follow-up times over the full study window. The average follow-up time to the third LDL measure was three years and the maximum was ten. For this analysis, we redefined *PDC_k_* as the average PDC value between time point *k-1* and *k*. For example, if k = 1, then k-1 indexes the baseline LDL and k indexes the first follow-up LDL. We assumed that *PDC_1_* up to *PDC_k_* exerted additive linear effects on *LR_k_*. Our goal was to estimate these effects, averaged over the study population, by adjusting for fixed and time-varying confounding. The causal diagram in Figure 4 Panel (a) shows the assumed data structure for our analysis.

**Figure 3.**
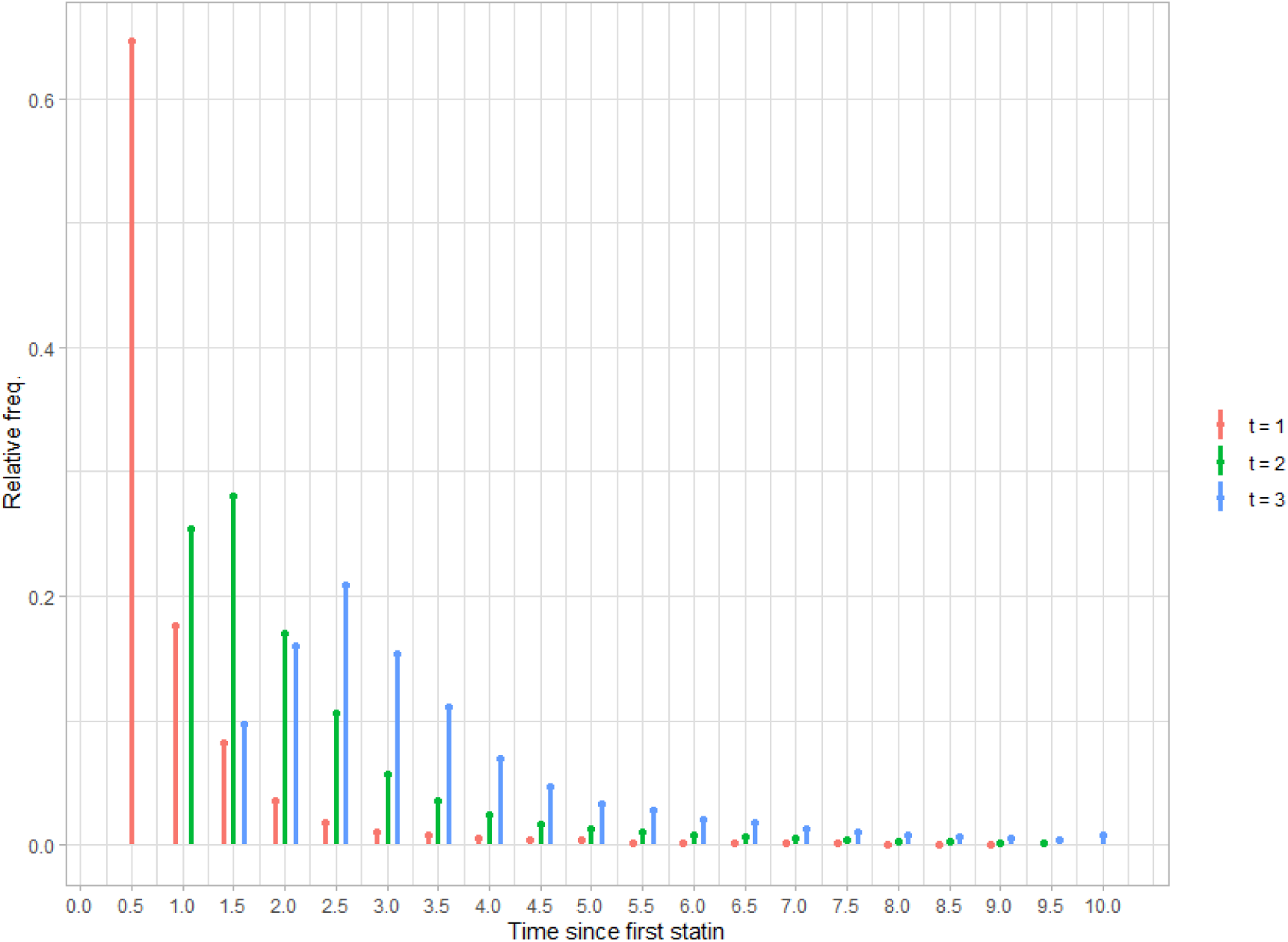
Frequency distribution of first three post-statin LDL measures times. The x-axis shows the time length by year, and y-axis shows the relative frequency

**Figure 4.**
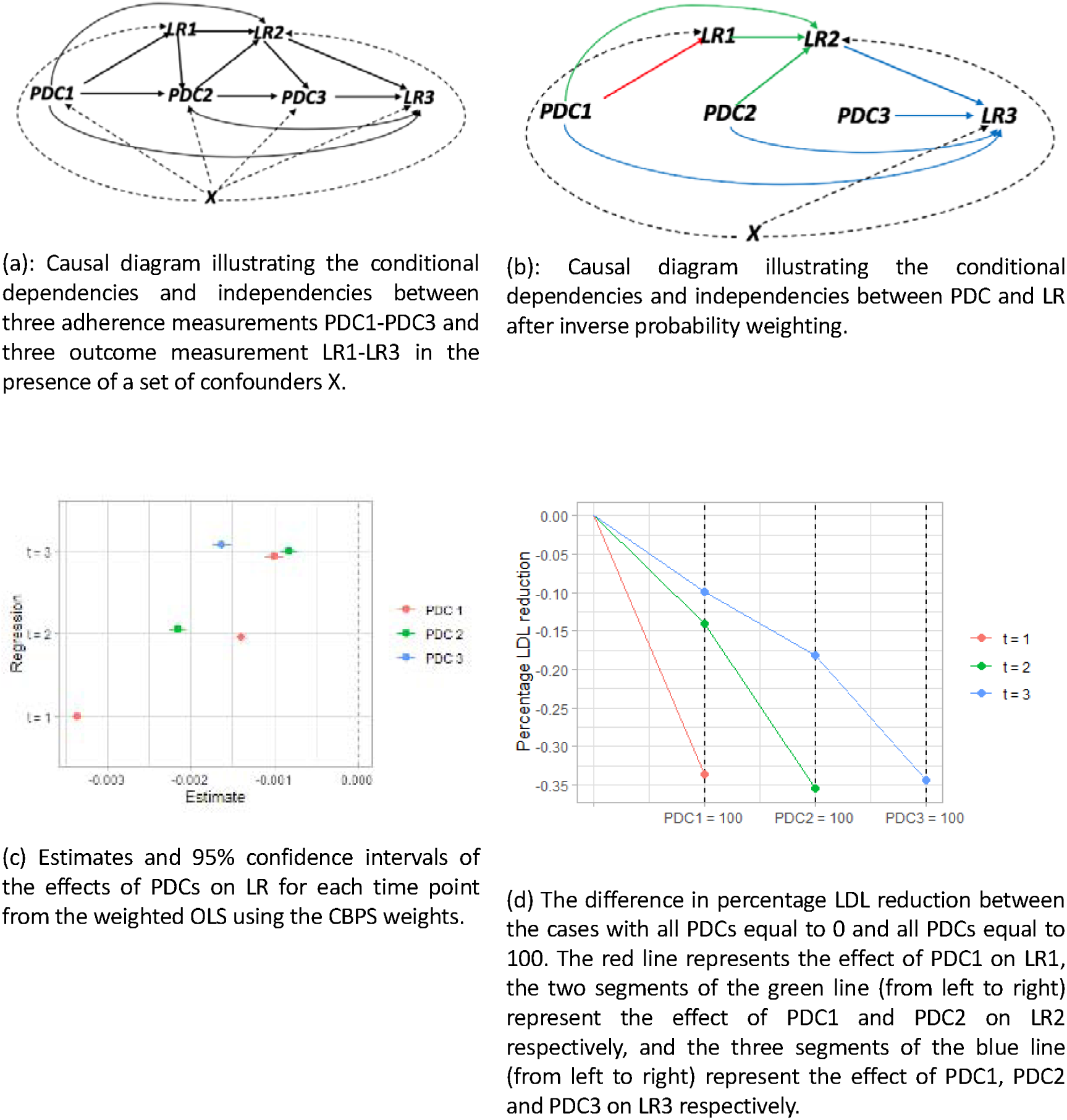
Causal diagrams illustrating the model set up of and results of the inverse probability weighted analysis.

We made the critical assumption that all the confounders of PDC and LR were observable and could be controlled for in the analysis. We include age at the first statin prescription, sex, pre-statin baseline LDL (i.e., *LDL_O_* ), education and assessment centre as fixed confounding factors. Previous PDC and LR were assumed to act as time varying confounders of future PDC and LR measures. We also included patient visit time for each post-statin LDL measure as the time-varying confounder. We utilized the inverse probability weighting (IPW) method, which is a two-step procedure for causal estimation under the observable confounding assumption. In stage 1, we create a pseudo-population by assigning everyone in the sample a weight so that, in this weighted pseudo-population, the exposure PDC is balanced across all the confounding covariates (see Figure 4 Panel (b) for illustration). We applied the Covariate Balancing Propensity Score (CBPS) method (26) to estimate the weight for each individual, using both the parametric and non-parametric (npCBPS) options. After obtaining the weights, we evaluated the covariate balance in the pseudo-population by calculating the weighted Pearson and Spearman correlation between each exposure and confounding covariate. Following the practice in (27), if the size of the correlation fell below the cut-off value 0.1, then the corresponding covariate was deemed sufficiently balanced. In stage 2, for each time point k, we estimated the effect of each *PDC_1_* to *PDC_k_* on *LR_k_* by the weighted OLS regression (regressing *LR_k_* on *PDC_1_* to *PDC_k_* simultaneously) using the weights obtained from the first step. The effect of the contemporaneous *PDC_k_* on *LR_k_* is the direct effect, while the effect of intervening on a previous PDC on *LR_k_* incorporates both the direct effect and the indirect effect via the earlier LR measures. If a covariate failed the balancing test, we included it as a covariate in the regression (28). See the Appendix for further technical details on the causal modelling and estimation methods.

We conducted LDL analyses (Analysis 1 and Analysis 2) separately for patient groups with and without prevalent CVD.

### Genome Wide Association Studies (GWAS)

We performed GWAS analysis of PDC traits using REGENIE v3.3(29) adjusted for age, sex, assessment centres, and genotyping microarray (2 categories) to identify genetic variants predicting PDC. REGENIE adjusts for sample relatedness and population structure during analysis. . We studied 16million genetic variants from the imputed data released by UKB (category 100319, (30)), where the imputation quality was >30% and Minor Allele Frequencies were >0.1%. P-values < 5*10^-8^ were taken as genome-wide significant.

### Time-to-event analysis of long-term clinical outcomes

We conducted a time-to-event analysis beginning 3 months post-statin initiation. Patients exited the model at the event date or end of follow-up. The model adjusted for sex, age at first statin prescription, education level, assessment centre, and baseline binary event diagnosis. Adherence levels were categorized as ≤50%, 50–95%, and >95% and used as exposure.

Unless otherwise stated all analyses were performed using R v4.3.1 and the package ‘survival’ (v3.5- 8) was used for time-to-event analysis. Package ‘CBPS’ (version: 0.23) was used for longitudinal causal analyses.

## Results

### LDL

Of the 76,909 UKB participants ever prescribed statins in their available primary care data (33% of all participants with primary care data), 41,316 had LDL measurements prior to statin initiation (<180 days pre-statin; Mean: 32, SD: 38, Median: 16) and within 1 year of first statin prescription (post- statin LDL measurement; Mean: 123 days, SD: 96, Median: 92). Of these, 37,297 were of European- like genetic ancestry (Figure 1). Mean age at statin initiation was 61 years (SD 7.3, range 40-79), and 44.4% were female (Table 1).

### Analysis 1: Cross-sectional analysis of LDL change

The mean pre- and post-statin LDL was 3.8 mmol/L (SD 1.05) and 2.5 mmol/L (SD 0.95) respectively. Individuals with pre-statin CVD (secondary prevention group) had, on average, baseline LDL levels 0.59 units lower (95% CI 0.62 to 0.56, p=7.6x10-355) and pre-statin CVD was associated with a 0.12 unit increase in dose intensity (95% CI 0.1 to 0.13, p=3.7x10-71) than those without pre-statin CVD. Across all individuals, we found a U-shaped relationship between LDL-c reduction and the time to first follow-up, with a maximum reduction for those returning for follow-up between weeks 5-8 of 26% (95%CI 23-29) (Appendix - Figure 1). The mean *PDC_1_* value was 94.7% (IQ 85.5-105) and the prevalence of PDCs is as follows: PDC = 50% (8%), PDC = 50-95% (33%), and PDC > 95% (59%). Stratifying the analysis by Individuals who had <=95% and >95 *PDC_1_* revealed the U-shape was driven by the former patient group rather than the latter (Appendix - Figure 2).

In the primary prevention group, the high PDC group, which was also more adherent to post-statin LDL follow-up (Figure 5-A), achieved the greatest LDL reduction, with a 40% reduction (LDL ratio ∼0.6) by 15 weeks. In the secondary prevention group, post-statin LDL levels were reduced to below 2 mmol/L, as recommended by NICE guidelines, in 31% of individuals. Those in the high PDC group (>95%) showed the greatest LDL reduction by 15 weeks, with 57% achieving LDL ≤2, compared to 23% in the low PDC group (p=2x10⁻³¹). The group sizes and densities indicate greater adherence to LDL check-ups in the high PDC group, which was associated with the most significant LDL reduction over time (Figure 5-B).

**Figure 5.**
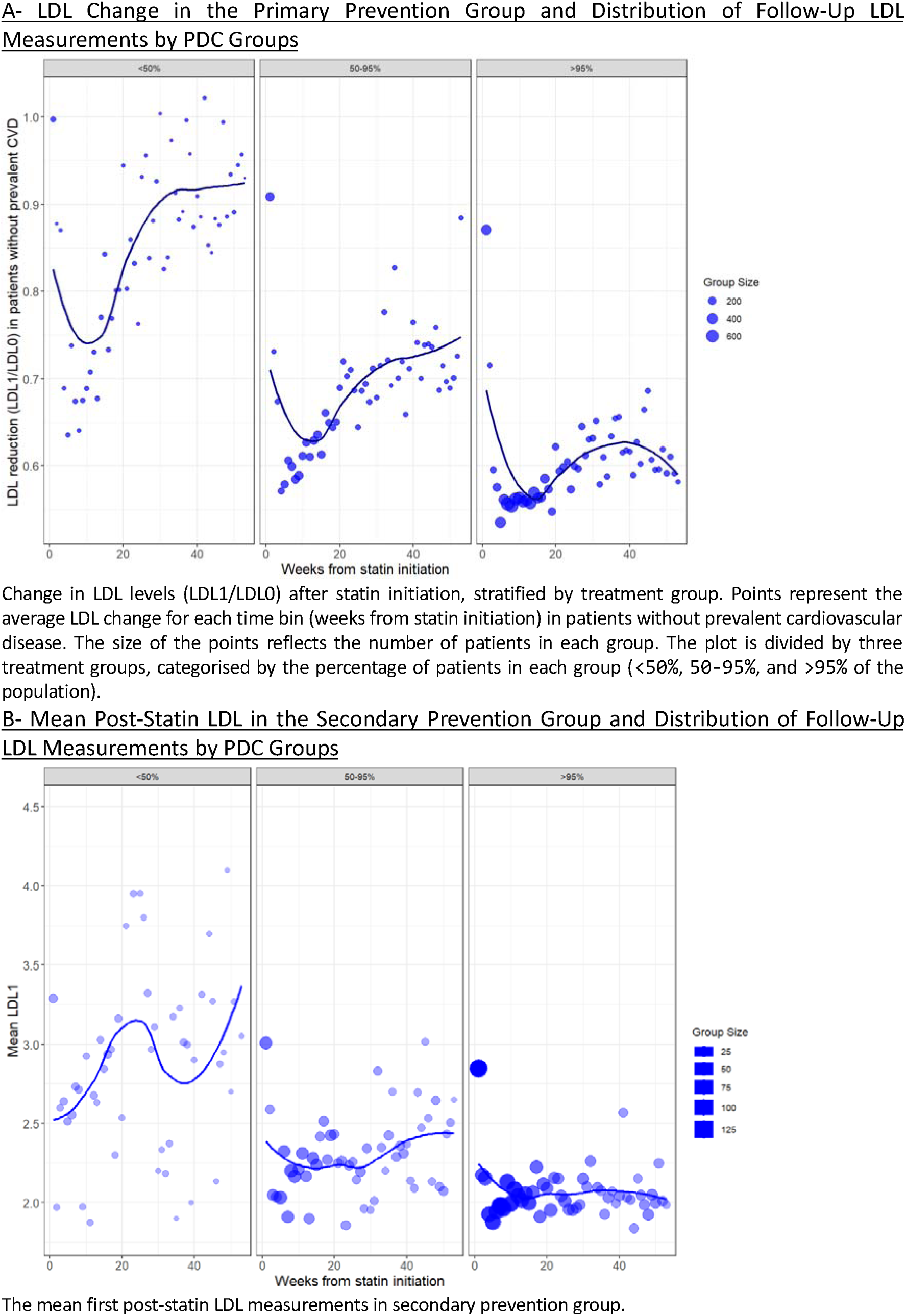
LDL Change in Primary Prevention & Mean LDL in Secondary Prevention Groups.

Finally, SLCO1B1*5 genotype was seen to alter the relationship between follow-up time and LDL reduction: the homozygotes (i.e., rs4149046 CC genotype) had the smallest LDL-c reduction in the quadratic model in year 1 (Appendix - Figure 3). The polygenic score for Alzheimer’s disease was associated with a reduced LDL reduction (Coef: 4.5×10⁻³, 95% CI 1.7×10⁻³ to 7.3×10⁻³, p=0.002). Supplementary Table 4 for the other polygenic scores.

### Analysis 2: Longitudinal analysis on LDL trajectories

Using the weights obtained from the stage 1 weighting procedure, we examined the weighted pair-wise correlations between *PDC_k_* and each of its constituent variables with both the Pearson and Spearman correlation. All covariates except for the patient visit times had average correlations (both Pearson and Spearman) with *PDC_k_* that fell below the 0.1 threshold (Supplementary Tables 5-6). We therefore explicitly included patient visit times as covariates in the stage two weighted regressions.

Model-derived estimates of the effects of PDCs on LR with the 95% confidence intervals using the CBPS weights are shown in Figure 4 Panel (c). All three PDC measures have significant (p < 0.05) negative effects on LDL reduction (Supplementary Table 7). Firstly, for all three time points, the most recent PDC measure, *PDC_k_* , had the largest estimated effect on *LR_k_*. Secondly, the initial adherence level PDC_1_ had a larger estimated impact on LR_3_ than PDC_2_. Thirdly, while PDCs at different time points had different estimated effects, the total effect estimate (expressed as a summation across the time points) remained remarkably constant. For illustration, we calculated the average difference in LDL reduction if all individuals’ PDCs were set to 100 versus if all PDCs were set to 0 in Figure 4 Panel (d). This revealed that LDL percentage reductions of approximately 35% can be achieved for sustained, full adherence to statins. Another relevant measure for each patient is the LDL reduction if they could maintain their own personal maximum PDC level across each time point compared to their observed PDC level at each time point. Our model suggests this difference would be approximately 6%. We conducted the analysis using the non-parametric option of the CBPS method (npCBPS) as well, with consistent results (Supplementary Table 8).

The longitudinal analysis of LDL changes with sustained PDC intervention in the primary prevention subgroup (n = 10,971) yielded results consistent with those in the main analysis of the full sample (Supplementary Tables 9-10). There was insufficient statistical power for analysis of the secondary prevention subgroup due to small sample size (n = 2,344).

### Predictors of PDC_1_

Male participants (Coef -0.64, 95% CI -1.25 to -0.04, p=0.03) and current smokers (Coef -3.66, 95% CI -4.61 to -2.40, p=7x10^-14^) were less adherent (lower PDC). Participants with higher pre-statin LDL-c, higher educational attainment, more prevalent diseases, and who were older at first stain prescription were more adherent (higher PDC). BMI was not associated with PDC (Coef 0.01, 95% CI - 0.05 to 0.07). Genetic predictors of reduced PDC included liability to schizophrenia (Coef_top 20%_ -1.94, 95%CI -2.69 to -1.19), whilst genetic liability to cardiovascular diseases increased PDC (Coef_top 20%_ 1.30, 95%CI 0.55 to 2.05) (Supplementary table 11). rs4149046 CC (*SLCO1B1*\*5), was not significantly associated with PDC in linear regression models (Coef -0.002, 95% CI -1.56 to 1.55, p=0.1). See table 2 for the details.

**Table 2.** Associations between risk factors and PDC (proportion of days covered) as adherence measure using linear regression in GP-prescribed statin patients in UK Biobank.

| Exposure | n | Coef | 95% | CI | p |
| --- | --- | --- | --- | --- | --- |
| **Observational<sup>^</sup>** |  |  |  |  |  |
| Sex (male) |  | -0.64 | -1.25 | -0.04 | 3.7E-02 |
| Age at first statin |  | 0.34 | 0.30 | 0.39 | 5.8E-54 |
| BMI |  | 0.01 | -0.05 | 0.07 | 7.5E-01 |
| LDL pre statin (GP) |  | 0.55 | 0.30 | 0.80 | 1.3E-05 |
| Smoking |  |  |  |  |  |
| Never smoked | 24932 | ref | - | - | - |
| Ex-smoker | 22982 | 0.63 | -0.003 | 1.26 | 5.1E-02 |
| Current smoker | 7014 | -3.66 | -4.61 | -2.70 | 7.2E-14 |
| Education |  |  |  |  |  |
| None | 15583 | ref | - | - | - |
| CSEs | 1584 | 0.17 | -1.63 | 1.98 | 8.5E-01 |
| GCSEs/O-levels | 6729 | 1.38 | 0.38 | 2.38 | 6.9E-03 |
| A-levels/NVQ/HND/HNC | 9403 | 1.24 | 0.32 | 2.15 | 7.9E-03 |
| Prof.qual | 7952 | 1.92 | 0.98 | 2.87 | 6.5E-05 |
| Degree | 13110 | 2.52 | 1.69 | 3.34 | 2.4E-09 |
| **Multimorbidity\*** |  |  |  |  |  |
| No add. Disease | 3127 | ref | - | - | - |
| 1 Disease | 7644 | 2.85 | 1.33 | 4.37 | 2.4E-04 |
| 2 Diseases | 9128 | 3.97 | 2.49 | 5.45 | 1.5E-07 |
| 3 Diseases | 8584 | 3.87 | 2.38 | 5.36 | 3.7E-07 |
| 4 Diseases | 7087 | 4.30 | 2.78 | 5.82 | 3.2E-08 |
| 4+ Diseases | 19743 | 4.44 | 3.05 | 5.84 | 4.4E-10 |
| **Polygenic Scores<sup>`</sup>** |  |  |  |  | **p<sub>BH</sub>** |
| SCZ | 49873 | -0.58 | -0.81 | -0.34 | 5.9E-05 |
| CAD | 49873 | 0.49 | 0.25 | 0.72 | 8.4E-04 |
| CVD | 49873 | 0.44 | 0.20 | 0.67 | 3.4E-03 |
| **Genotype** |  |  |  |  | **p** |
| rs4149056 TT | 36079 | ref | - | - | - |
| rs4149056 TC | 12759 | 0.09 | -0.45 | 0.62 | 7.56E-01 |
| rs4149056 CC | 1162 | -0.002 | -1.56 | 1.55 | 9.98E-01 |

Positive estimates (Coef: Coefficient) have increased PDC.
<sup>^</sup> Age at first statin: age at the first prescription by GP
\*Multimorbidity includes 84 diseases including cardiovascular diseases. No add. Disease represents patients with no disease prior to their first statin prescription.
<sup>`</sup>Three of 36 polygenic scores derived by Genomics PLC tested are significant after The Benjamini-Hochberg multiple testing adjustments. The units are scaled; distribution has a zero mean and unit variance. SCZ: Schizophrenia, CAD: Coronary Arteria Disease, CVD: Cardiovascular Disease. Polygenic scores are continuous, and genotype are tested in European-like participants only to minimise population stratification effect.

### GWAS

We performed GWAS analysis of two PDC traits, using REGENIE to analyse 16million genetic variants with MAF>0.001 and INFO>0.3. One genetic variant was significantly (p<5*10^-8^) associated with “PDC until LDL1” in linear regression models: rs548267220 (C>G, MAF=0.006, INFO=0.8, BETA=12.7, p=4.8*10^-8^), intronic for gene *RUNX3*. Two genetic variants were significantly (p<5*10^-8^) associated with “PDC >95% vs. <=95%” in logistic regression models: rs9439705 (G>A, MAF=0.92, INFO=0.99, BETA=-0.13, p=1.4*10^-8^), located 30kbp from *KLHDC7A*, and rs75103961 (C>T, MAF=0.002, INFO=0.92, BETA=-0.77, p=4.0*10^-8^), intergenic on chromosome 12. For details see Supplementary Tables 12.

rs548267220 has not appeared in the GWAS catalog previously (5^th^ Nov 2024), though there are 142 entries for *RUNX3* including haematological traits, kidney function, allergic diseases, blood pressure, and obesity. Neither rs9439705 nor rs75103961 have appeared in the GWAS catalog previously (5^th^ Nov 2024). *KLHDC7A*, encoding protein kelch domain containing 7A, has previously been linked to LDL cholesterol, kidney function, and other traits in the GWAS catalog.

### Time-to-event analysis

We analysed up to 55,784 patients across three PDC groups (low: <50%, moderate: 50-95%, high: >95%). Hypertension was the most prevalent disease (43%). Key differences between PDC groups included CHD (19% in low PDC, 17% in high PDC), cataract (8% in low PDC, 10% in high PDC), and chronic kidney disease (8.5% in low PDC, 10% in high PDC) (Supplementary Table 13).

Patients with high PDC had a decreased risk of ischemic stroke (HR 0.67, 95% CI 0.54-0.83, p=10^-2^), coronary heart disease (HR 0.78, 95% CI 0.73-0.84, p=2x10^-8^), transient ischemic attack (HR 0.79, 95% CI 0.68-0.92, p=4x10^-2^); but an increased risk of cataract (HR 1.20, 95% CI 1.07-1.34, p=4x10^-2^), iron deficiency anaemia (HR 1.3, 95% CI 1.09-1.54, p=5x10^-2^), chronic kidney disease (HR 1,19, 95% CI 1.06-1.32, p=4x10^-2^) and enthesopathy of the upper limbs (HR 1.16, 95% CI 1.06-1.26, p= 4x10^-2^) compared to those with low PDC (Table 3). P-values were adjusted using the Benjamini-Hochberg correction method.

**Table 3.** Associations between future diseases and PDC (low/high) PDC>95 versus PDC<50.

| Outcome | % high PDC<br>(low PDC) | n case<br>high PDC | n case<br>total | HR | 95% | CI | p_bh |
| --- | --- | --- | --- | --- | --- | --- | --- |
| **CHD** | 16.62 (18.56) | 5455 | 9421 | 0.78 | 0.73 | 0.84 | 2.E-08 |
| **Isch Stroke** | 1.56 (2.30) | 513 | 928 | 0.67 | 0.54 | 0.83 | 1.E-02 |
| **Cataract** | 9.68 (7.81) | 3177 | 5237 | 1.20 | 1.07 | 1.34 | 4.E-02 |
| **CKD** | 10.04 (8.57) | 3297 | 5370 | 1.19 | 1.06 | 1.32 | 4.E-02 |
| **Enthesopathy U.** | 12.62 (12.66) | 4144 | 7162 | 1.16 | 1.06 | 1.26 | 4.E-02 |
| **TIA** | 4.00 (4.73) | 745 | 2221 | 0.79 | 0.68 | 0.92 | 4.E-02 |
| **IDA** | 3.93 (3.38) | 1289 | 2145 | 1.30 | 1.09 | 1.54 | 5.E-02 |

We performed a time-to-event analysis in statin patients, starting 3 months post-initiation.
Patients exited the model at either the event date or the end of follow-up. The model
adjusted for sex, age at first statin use, education, assessment center, and prevalent
diagnosis of the event (binary). We compared adherence levels ( $\leq 50\%$ , $50-95\%$ , and $>95\%$ ),
showing here on results for the high adherence group (PDC $>95\%$ ).
Outcomes were derived from General Practice (GP) and hospital records. Abbreviations: CHD
(Coronary Heart Disease), Isch (Ischemic), CKD (Chronic Kidney Disease), Enthesopathy U
(Upper), TIA (Transient Ischemic Attack), IDA (Iron Deficiency Anemia), % (prevalence), n
(total number of cases), HR (Hazard Ratio), p\_bh (Benjamini–Hochberg corrected p-value).

## Discussion

Using real-world data, we identify substantial variability in the time to first on-treatment LDL and adherence to statin medication, highlighting a gap between NHS guidelines and the actual level of achieved treatment response monitoring. Our work highlights important predictors of reduced statin effectiveness, such as pharmacogenetic variants, polygenic scores, but most of all, sustained adherence to statin medications. Higher adherence to treatment was associated with greater adherence to therapy overall, leading to more significant LDL reductions over time. This suggests that improved service provision and patients’ factors—such as increased statin use, better understanding of treatment by patients, more frequent GP visits for LDL checks, and tailored treatments addressing low-adherence risk factors—could enhance efficacy, safety, and long-term patient outcomes.

When quantifying adherence using the Proportion of Days Covered (PDC) measure, we assumed that patients adhered to their prescribed medication regimens. However, the UKB lacks detailed data on actual medication intake and GP dosage instructions (e.g., whether a patient is taking 2x40 mg versus 1x80 mg). Therefore, we restricted our PDC analysis to patients with a PDC of less than 200%, under the assumption that they might be taking two tablets per day as prescribed. As a result, our adherence rate is higher than in previous Finnish cohorts, likely due to their smaller PDC cut-off and the fact that 96% of statin users in Finland were prescribed one tablet daily (31,32). Despite these differences, the extent to which LDL change is strongly predicted by our PDC measure indicates that it is capturing the underlying relationship between medication adherence and effectiveness of treatment.

To add further weight to this, our finding of an increased risk of iron deficiency anemia (IDA) in patients with higher statin adherence (HR 1.30: 95% CI 1.12–1.70) is consistent with previous research: a hypothesis-free analysis of a Korean cohort (7,847 statin users vs. 39,235 non-users) similarly identified a higher risk of IDA (95% CI 2.11–12.03) linked to statin use among over 100 tested diseases, supported by Mendelian Randomisation analysis (33). The possible mechanism between iron and statins can be linked to statin’s capacity to favourably modulate iron homeostasis (34), which might be beneficial for CVD risk as abnormal iron accumulation in cells increases oxidative stress, vascular inflammation, and the development of atherosclerosis (34,35). A review of over 313,200 patients (36) found a moderate increase in cataract risk with statin use in cohort studies, which had long follow-up periods (∼ >5 years) and various baseline characteristics, while case-control studies and randomized trials showed no significant risk. Although our findings support previous cohort studies with long-term follow-up, the review concludes that there is likely no significant link between statin treatment and cataracts, emphasizing that the benefits of statin therapy outweigh potential risks.

In exploratory analysis to evaluate the effect of a sustained PDC intervention on LDL reduction using longitudinal causal models, we made a critical assumption that all confounders between PDC and LDL across three time points were observable and could be controlled for in the analysis using inverse probability weighting. The resulting estimates suggest that approximately 35% reductions in LDL are eminently achievable in the UKB population under full adherence. Ensuring high adherence in the most recent period was estimated to have, unsurprisingly, the largest influence on patients’ LDL reduction, but ensuring high adherence in their initial treatment phase was interestingly the next most effective way to achieve this. This again highlights the importance of early follow-up monitoring after statin initiation. A possible direction for future research is to utilize pharmacogenetic predictors of treatment adherence within a Mendelian randomization framework (37,38), to facilitate this analysis under a relaxation of the observable confounder assumption. Unfortunately, we did not identify any suitably strong genetic predictors of PDC in the UKB primary data reaching genome wide significance. This is consistent with findings from the FinnGen/EstoniaBiobank study (18) , which also failed to identify any genome-wide significant genetic variants associated with PDC in statin users (18). Future work will expand this search to include other cohort studies and to consider other measures of adherence beyond PDC.

A PDC below 50% is particularly important, as it indicates that even if patients are taking 1/2 tablets daily, their adherence is still insufficient to fully meet the prescribed regimen. Consequently, patients with low adherence had a 27% higher risk of coronary heart disease and a 50% increased risk of ischemic stroke compared to those with high adherence. Understanding and improving adherence is crucial to reducing cardiovascular events. Predictors of low adherence to statins included genetic liability to schizophrenia. Likewise, a study from the Estonian Bank (39) highlighted that genetic liability to schizophrenia was associated with schizophrenia diagnoses in this cohort. For these patients, higher adherence to antipsychotic medications reduced metabolic syndromes, which are known to elevate the risk of heart disease and stroke. Our research suggests that this could be due indirectly to higher levels of statin adherence rather than a direct effect of antipsychotic medication.

When investigating genetic predictors of on-statin LDL response, we initially focused on the pharmacogenetic polygenic risk score reported by Mayerhofer et al (40). We did not find the score to be predictive of an LDL reduction as a whole. rs2900478, a single SNP included in the polygenic score, was highly correlated (r^2^=0.96) with the well-known statin pharmacogenetic variant SLCO1B1*5 (rs4149056). We suspected that the score effect could be attributed to the statin transporter gene SLCO1B1, and we continued our analysis focusing on this gene alone. Consistent with our previous work, where we found that SLCO1B1*5 was associated with discontinuation of statin therapy (41), we here report that *5 is associated with attenuated LDL reduction following statin initiation. We found no association between SLCO1B1*5 and PDC (adherence) perhaps because, by definition, those with multiple prescriptions meeting inclusion criteria have not yet discontinued. When taken together, our analyses support the known pharmacogenetic effect of *5 on statin effectiveness, and potential impact on adverse events.

In conclusion, this is the first study, to our knowledge, that uses a large cohort with linked genetics and primary care health records to examine the effect of adherence on LDL change and long-term outcomes, and identify risk factors for adherence using genetic and causal inference models. Our work emphasizes insufficient LDL control with a need for LDL monitoring in line with clinical guidelines, and a clear clinical benefit of adhering to prescribing guidance, which reinforces the importance of patient education on statin indication and adherence.

## Data Availability

The data that support the findings of this study are available from UK Biobank but restrictions apply to the availability of these data, which were used under license for the current study, and so are not publicly available. Data are however available from the authors upon reasonable request and with permission of UK Biobank (www.ukbiobank.ac.uk/register-apply)

## Declarations

### Ethics approval and consent to participate

The collection and use of UK Biobank data was approved by the North West Multi-Centre Research Ethics Committee (Research Ethics Committee reference 11/NW/0382).

## Consent for publication

UK Biobank participants gave informed consent for their data to be used in health research.

## Availability of data and materials

The data that support the findings of this study are available from UK Biobank but restrictions apply to the availability of these data, which were used under license for the current study, and so are not publicly available. Data are however available from the authors upon reasonable request and with permission of UK Biobank (www.ukbiobank.ac.uk/register-apply). GWAS summary statistics will be posted to Zenodo and a DOI provided following acceptance.

## Authors’ contributions

DT and XL generated data, performed analyses, interpreted results, created the figures, searched the literature, and co-wrote the manuscript. JM and DG provided expert clinical interpretation of the data and contributed to the manuscript. LP interpreted results, searched the literature, and co-wrote the manuscript. JB oversaw interpretation, contributed to data analysis, and co-write the manuscript.

## Acknowledgements

Access to UK Biobank was granted under Application Number 14631. The authors would like to acknowledge the use of the University of Exeter High-Performance Computing (HPC) facility in carrying out this work. For the purpose of open access, the author has applied a ‘Creative Commons Attribution (CC BY)’ licence to any Author Accepted Manuscript version arising from this submission.

## Appendix

### Methods

#### Cross sectional modelling of LDL reduction explained by week of follow up

We tested associations between LDL change and the time interval between their pre- and post- treatment measurements, using a linear regression model, adjusted additionally for dose group of first atorvastatin and simvastatin prescribed in year 1. Specifically:

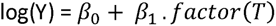

Y = LDL change (LDL1/LDL0)

T = Time interval between their pre- and post-treatment measurements

We organised doses into a categorical variable based on NICE guidelines (21) for dose equivalence, as shown below:

**Table 1.** Dose intensity classification based on the NICE Guideline.

| Drug | 10 mg | 20 mg | 40 mg | 80 mg |
| --- | --- | --- | --- | --- |
| Simvastatin | low | middle | Middle | High |
| Atorvastatin | middle | high | High | High |

**Table 2.** Prevalence of the dose intensity groups in individuals prescribed statins in the UK Biobank GP data.

| Dose intensity | N total | % | N (in both LDL0 and LDL1 available) | % |
| --- | --- | --- | --- | --- |
| low | 5,808 | 7.7 | 2,355 | 6.4 |
| middle | 57,293 | 76.3 | 29,560 | 80.4 |
| high | 12,013 | 16.0 | 4,833 | 13.2 |
| total: | 75114 |  | 36,748 |  |

We repeated this analysis in *PDC*_1_ groups (<=95% and >95%).

#### Modelling LDL reduction by genotype, PDC and time of follow up measure

We tested associations between LDL change and rs4149056 using a linear model that allowed for a quadratic effect of time to follow up on treatment in European-like participants and an interaction between time on treatment and rs4149056:

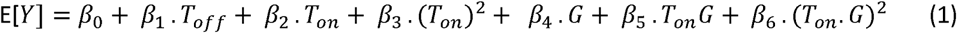

*Y* = *LDL change* (LDL1/LDL0)

*T_Dff_* = Time between pre-statin LDL measurement and the statin initiation

*T_on_*= Time to first LDL measures after the statin initiation x (dose group).

*G* = rs4149056 (number of copies)

Model (1) additionally adjusted for 10 genetic principal components (PCs).

Next, we tested the association between LDL change and *PDC*_1_ using model (1), but with PDC in place of the G variable, rs4149056. This analysis did not adjust for genetic PCs but accounted for the prevalence of CVD before statin use, including abdominal aortic aneurysm, atrial fibrillation, coronary heart disease, peripheral arterial disease, all types of stroke, and thromboembolic disease.

#### Causal modelling of hypothetical LDL reduction via sustained PDC intervention

Let the vector 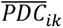 denote the adherence history of patient *i* up to time point *k* so that: 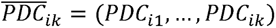 ^-^We define 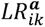 as the potential LDL reduction at time point *k* given that 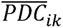 takes the value *a*. We assume that *PDC_i_*_1_ to *PDC*_ik_ exert linear additive effects on *LR_ik_*.

More concretely, for each time point k, we define the estimand of interest as the contrast between the potential outcomes under PDC history profile ***a***.+**1** and ***a***., which we assume obeys the following parametric relationship: 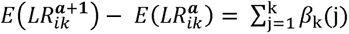

where *k* = 1,2,3 and *β*_k_(j) represents the causal effect of *PDC_i_*_1_ on *LR_ik_* with *j* = 1, …, *k*. We aim to estimate *β*_k_(j) with the inverse probability weighting (IPW) method under the assumption that all the confounders between PDC and LR are observable and can be controlled for in the analysis. Since we have a continuous exposure variable PDC, according to (20) and (26), the weight for individual *i* at time point *k* is given by

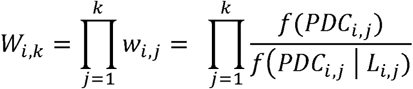

with *k* = 1, 2,3. *f*(*PDC_i,j_*) and *f*(*PDC_i,k_* | *L_i,j_*) are the marginal and conditional density functions respectively. *PDC_i,j_* is the observed value of the PDC for individual *i* at time point *j* and *L_i,j_* summarizes all fixed covariates mentioned in the main text, the previous exposure *PDC_i,j-l_* and previous outcome *LR_i,j-l_* (for 1= 2, 3), and the patient visit time, denoted by *t_i,j_*.

We estimate each *w_i,j_* (*j* = 1,2, 3) with the Covariate Balancing Propensity Score (CBPS) method proposed by (26). The method was implemented with the R functions ‘CBPS’ and ‘npCBPS’ (for the non-parametric option) from the R package ‘CBPS’ (https://cran.r-project.org/web/packages/CBPS/index.html). We ran the functions with default settings except that we set ‘method = exact’ for CBPS. For each *j* = 1, 2,3, we estimated *w_i,j_* by fitting the model of *PDC_i,j_* ∼ *L_i,j_*. For a given time point *k* (*k* = 1,2,3), *w_i,k_* is then obtained with 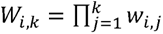_j_. We multiplied each *w_i,k_* by 10^l3^ for the second stage weighted OLS to avoid zero truncation due to machine tolerance issues. In the second-stage regression, we omit the intercept.

### Results

**Figure 1.**
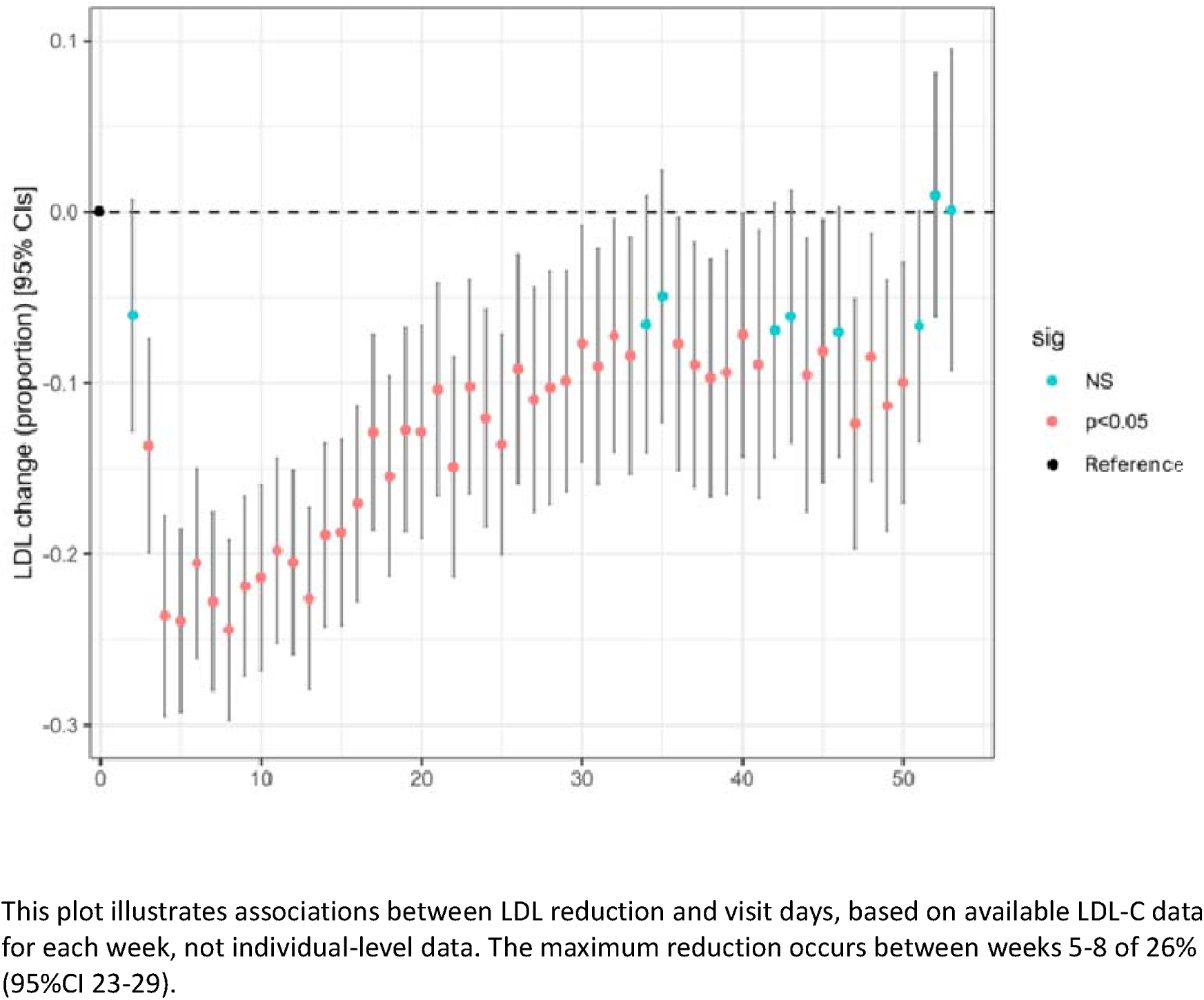
Associations between time to first GP LDL measure following statin initiation and LDL change.

**Figure 2.**
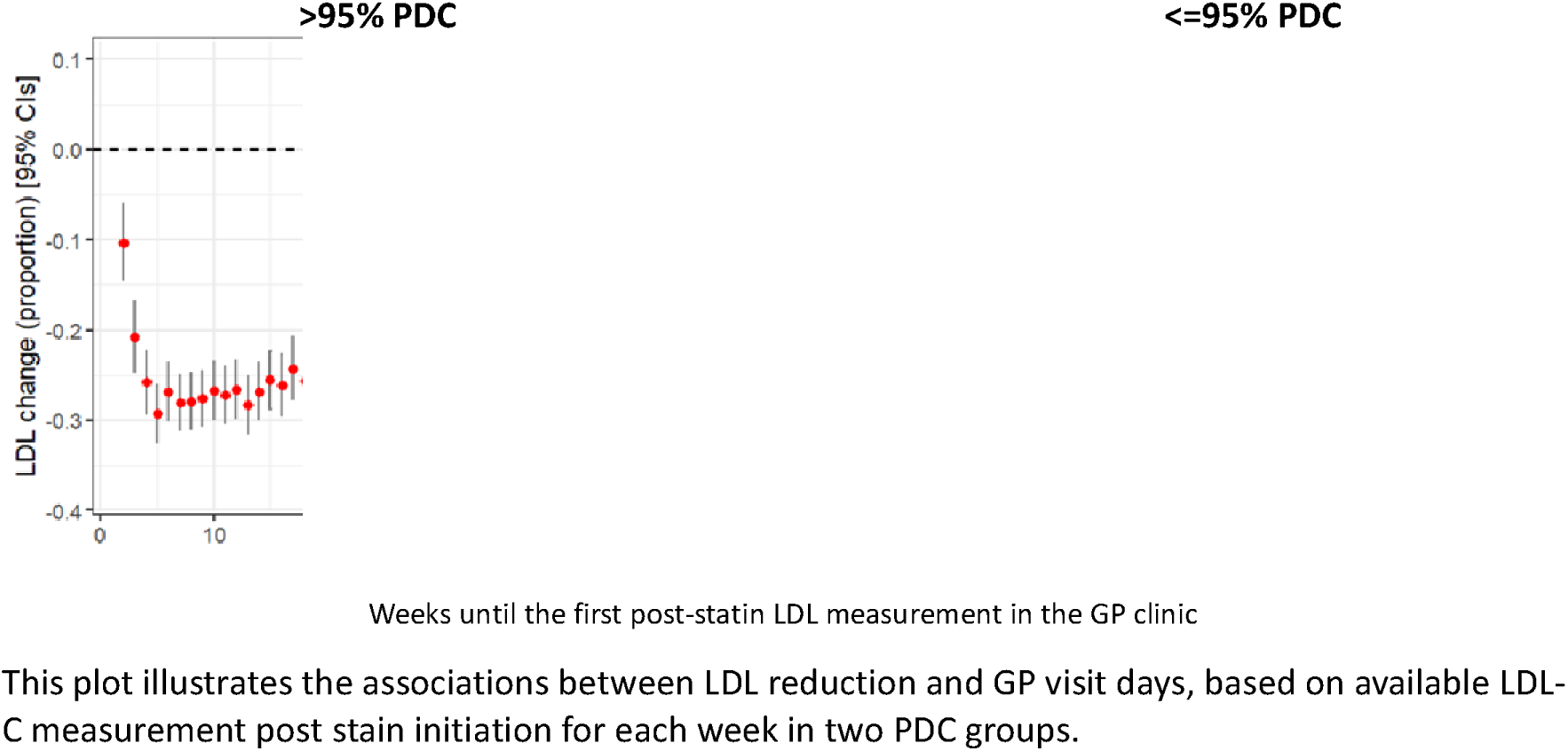
Associations between time to first GP LDL measure following statin initiation and LDL change by PDC groups.

**Figure 3.**
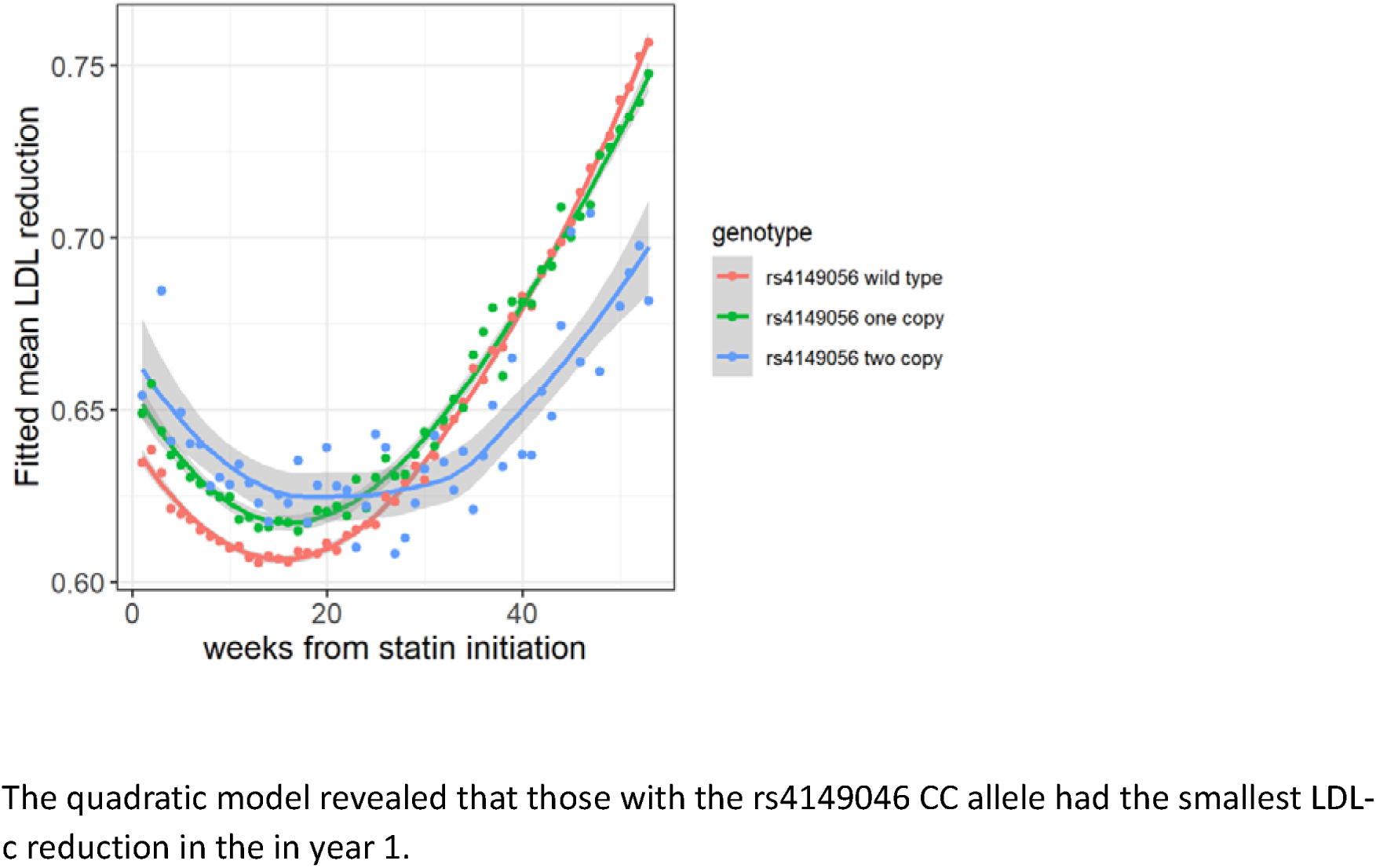
Quadratic model of LDL reduction estimated by SLCO1B1 rs4149046 genotype.

## REFERENCES

1. Roth GA, Mensah GA, Johnson CO, Addolorato G, Ammirati E, Baddour LM, et al. Global Burden of Cardiovascular Diseases and Risk Factors, 1990–2019. J Am Coll Cardiol. 2020 Dec;76(25):2982–3021.

2. Efficacy and safety of cholesterol-lowering treatment: prospective meta-analysis of data from 90,056 participants in 14 randomised trials of statins - PubMed [Internet]. [cited 2024 Oct 22]. Available from: https://pubmed.ncbi.nlm.nih.gov/16214597/

3. Recommendations | Cardiovascular disease: risk assessment and reduction, including lipid modification | Guidance | NICE [Internet]. NICE; 2023 [cited 2024 Jun 24]. Available from: https://www.nice.org.uk/guidance/ng238/chapter/recommendations#assessing-response-to-treatment-5

4. Virani SS, Newby LK, Arnold SV, Bittner V, Brewer LC, Demeter SH, et al. 2023 AHA/ACC/ACCP/ASPC/NLA/PCNA Guideline for the Management of Patients With Chronic Coronary Disease: A Report of the American Heart Association/American College of Cardiology Joint Committee on Clinical Practice Guidelines. Circulation [Internet]. 2023 Aug 29 [cited 2024 Dec 10];148(9):e9–119. Available from: https://www.ahajournals.org/doi/10.1161/CIR.0000000000001168

5. Arnett DK, Blumenthal RS, Albert MA, Buroker AB, Goldberger ZD, Hahn EJ, et al. 2019 ACC/AHA Guideline on the Primary Prevention of Cardiovascular Disease: A Report of the American College of Cardiology/American Heart Association Task Force on Clinical Practice Guidelines. Circulation [Internet]. 2019 Sep 10 [cited 2024 Dec 10];140(11):e596–646. Available from: https://www.ahajournals.org/doi/10.1161/CIR.0000000000000678

6. Mach F, Baigent C, Catapano AL, Koskinas KC, Casula M, Badimon L, et al. 2019 ESC/EAS guidelines for the management of dyslipidaemias: Lipid modification to reduce cardiovascular risk. Atherosclerosis. 2019 Nov;290:140–205.

7. Lloyd-Jones DM, Morris PB, Ballantyne CM, Birtcher KK, Covington AM, DePalma SM, et al. 2022 ACC Expert Consensus Decision Pathway on the Role of Nonstatin Therapies for LDL-Cholesterol Lowering in the Management of Atherosclerotic Cardiovascular Disease Risk. J Am Coll Cardiol. 2022 Oct;80(14):1366–418.

8. Cholesterol Treatment Trialists’ (Ctt) Collaboration. Efficacy and safety of more intensive lowering of LDL cholesterol: a meta-analysis of data from 170 000 participants in 26 randomised trials. The Lancet. 2010 Nov;376(9753):1670–81.

9. Wilkinson MJ, Lepor NE, Michos ED. Evolving Management of Low-Density Lipoprotein Cholesterol: A Personalized Approach to Preventing Atherosclerotic Cardiovascular Disease Across the Risk Continuum. J Am Heart Assoc. 2023 Jun 6;12(11):e028892.

10. Maningat P, Gordon BR, Breslow JL. How Do We Improve Patient Compliance and Adherence to Long-Term Statin Therapy? Curr Atheroscler Rep [Internet]. 2013 Jan [cited 2024 Oct 21];15(1):291. Available from: https://pmc.ncbi.nlm.nih.gov/articles/PMC3534845/

11. Bosworth HB, Granger BB, Mendys P, Brindis R, Burkholder R, Czajkowski SM, et al. Medication adherence: A call for action. Am Heart J [Internet]. 2011 Sep [cited 2024 Oct 11];162(3):412–24. Available from: https://linkinghub.elsevier.com/retrieve/pii/S0002870311004753

12. Rodriguez F, Maron DJ, Knowles JW, Virani SS, Lin S, Heidenreich PA. Association of Statin Adherence With Mortality in Patients With Atherosclerotic Cardiovascular Disease. JAMA Cardiol. 2019 Mar 1;4(3):206.

13. Hirsh BJ, Smilowitz NR, Rosenson RS, Fuster V, Sperling LS. Utilization of and Adherence to Guideline-Recommended Lipid-Lowering Therapy After Acute Coronary Syndrome: Opportunities for Improvement. J Am Coll Cardiol [Internet]. 2015 Jul 14 [cited 2024 Oct 21];66(2):184–92. Available from: https://www.sciencedirect.com/science/article/pii/S0735109715024754

14. de Lemos JA, Blazing MA, Wiviott SD, Lewis EF, Fox KAA, White HD, et al. Early Intensive vs a Delayed Conservative Simvastatin Strategy in Patients With Acute Coronary SyndromesPhase Z of the A to Z Trial. JAMA [Internet]. 2004 Sep 15 [cited 2024 Oct 21];292(11):1307–16. Available from: 10.1001/jama.292.11.1307

15. Aubert RE, Yao J, Xia F, Garavaglia SB. Is there a relationship between early statin compliance and a reduction in healthcare utilization? Am J Manag Care. 2010 Jun;16(6):459–66.

16. Natarajan P, Young R, Stitziel NO, Padmanabhan S, Baber U, Mehran R, et al. Polygenic Risk Score Identifies Subgroup With Higher Burden of Atherosclerosis and Greater Relative Benefit From Statin Therapy in the Primary Prevention Setting. Circulation. 2017 May 30;135(22):2091–101.

17. Mega JL, Stitziel NO, Smith JG, Chasman DI, Caulfield MJ, Devlin JJ, et al. Genetic risk, coronary heart disease events, and the clinical benefit of statin therapy: an analysis of primary and secondary prevention trials. The Lancet. 2015 Jun;385(9984):2264–71.

18. Cordioli M, Corbetta A, Kariis HM, Jukarainen S, Vartiainen P, Kiiskinen T, et al. Socio- demographic and genetic risk factors for drug adherence and persistence across 5 common medication classes. Nat Commun [Internet]. 2024 Oct 23 [cited 2024 Dec 17];15(1):9156. Available from: https://www.nature.com/articles/s41467-024-53556-z

19. Sadler MC, Apostolov A, Cevallos C, Ribeiro DM, Altman RB, Kutalik Z. Leveraging large-scale biobank EHRs to enhance pharmacogenetics of cardiometabolic disease medications [Internet]. 2024 [cited 2024 Oct 17]. Available from: http://medrxiv.org/lookup/doi/10.1101/2024.04.06.24305415

20. Olarte Parra C, Daniel RM, Bartlett JW. Hypothetical Estimands in Clinical Trials: A Unification of Causal Inference and Missing Data Methods. Stat Biopharm Res [Internet]. 2023 Apr 3 [cited 2024 Oct 21];15(2):421–32. Available from: https://www.tandfonline.com/doi/full/10.1080/19466315.2022.2081599

21. Cardiovascular disease: risk assessment and reduction, including lipid modification. Cardiovasc Dis.

22. Sampson M, Ling C, Sun Q, Harb R, Ashmaig M, Warnick R, et al. A New Equation for Calculation of Low-Density Lipoprotein Cholesterol in Patients With Normolipidemia and/or Hypertriglyceridemia. JAMA Cardiol [Internet]. 2020 May 1 [cited 2024 Dec 6];5(5):540. Available from: https://jamanetwork.com/journals/jamacardiology/fullarticle/2761953

23. Casanova F, Tian Q, Atkins JL, Wood AR, Williamson D, Qian Y, et al. Iron and risk of dementia: Mendelian randomisation analysis in UK Biobank. J Med Genet [Internet]. 2024 Jan 8 [cited 2024 Oct 23];jmg-2023-109295. Available from: https://jmg.bmj.com/lookup/doi/10.1136/jmg-2023-109295

24. Thompson DJ, Wells D, Selzam S, Peneva I, Moore R, Sharp K, et al. UK Biobank release and systematic evaluation of optimised polygenic risk scores for 53 diseases and quantitative traits [Internet]. 2022 [cited 2024 Jun 24]. Available from: http://medrxiv.org/lookup/doi/10.1101/2022.06.16.22276246

25. Murrin O, Mounier N, Voller B, Tata L, Gallego-Moll C, Roso-Llorach A, et al. A systematic analysis of the contribution of genetics to multimorbidity and comparisons with primary care data [Internet]. 2024 [cited 2024 Jun 24]. Available from: http://medrxiv.org/lookup/doi/10.1101/2024.05.13.24307009

26. Fong C, Hazlett C, Imai K. Covariate balancing propensity score for a continuous treatment: Application to the efficacy of political advertisements. Ann Appl Stat [Internet]. 2018 Mar 1 [cited 2024 Oct 21];12(1). Available from: https://projecteuclid.org/journals/annals-of-applied-statistics/volume-12/issue-1/Covariate-balancing-propensity-score-for-a-continuous-treatment--Application/10.1214/17-AOAS1101.full

27. Austin PC. Assessing covariate balance when using the generalized propensity score with quantitative or continuous exposures. Stat Methods Med Res [Internet]. 2019 May [cited 2024 Oct 21];28(5):1365–77. Available from: https://journals.sagepub.com/doi/10.1177/0962280218756159

28. Bishop CD, Leite WL, Snyder PA. Using Propensity Score Weighting to Reduce Selection Bias in Large-Scale Data Sets. J Early Interv [Internet]. 2018 Dec [cited 2024 Oct 21];40(4):347–62. Available from: https://journals.sagepub.com/doi/10.1177/1053815118793430

29. Mbatchou J, Barnard L, Backman J, Marcketta A, Kosmicki JA, Ziyatdinov A, et al. Computationally efficient whole-genome regression for quantitative and binary traits. Nat Genet [Internet]. 2021 Jul [cited 2024 Aug 31];53(7):1097–103. Available from: https://www.nature.com/articles/s41588-021-00870-7

30. Bycroft C, Freeman C, Petkova D, Band G, Elliott LT, Sharp K, et al. The UK Biobank resource with deep phenotyping and genomic data. Nature [Internet]. 2018 Oct [cited 2022 Dec 12];562(7726):203–9. Available from: http://www.nature.com/articles/s41586-018-0579-z

31. Cordioli M, Corbetta A, Kariis HM, Jukarainen S, Vartiainen P, Kiiskinen T, et al. Socio- demographic and genetic risk factors for drug adherence and persistence: a retrospective nationwide and biobank study across 5 medication classes and 1 845 665 individuals [Internet]. 2023 [cited 2024 Aug 13]. Available from: http://medrxiv.org/lookup/doi/10.1101/2023.10.09.23296740

32. Aarnio EJ, Martikainen JA, Helin-Salmivaara A, Huupponen RK, Hartikainen JEK, Peura PK, et al. Register-based predictors of adherence among new statin users in Finland. J Clin Lipidol [Internet]. 2014 Jan [cited 2024 Nov 19];8(1):117–25. Available from: https://linkinghub.elsevier.com/retrieve/pii/S1933287413002973

33. Ahn J, Lee S, Won S. Possible link between statin and iron deficiency anemia: A South Korean nationwide population-based cohort study. Sci Adv [Internet]. 2023 Oct 27 [cited 2024 Oct 21];9(43):eadg6194. Available from: https://www.science.org/doi/10.1126/sciadv.adg6194

34. Mascitelli L, Goldstein MR. Might the beneficial effects of statin drugs be related to their action on iron metabolism? QJM [Internet]. 2012 Dec 1 [cited 2024 Oct 21];105(12):1225–9. Available from: https://academic.oup.com/qjmed/article-lookup/doi/10.1093/qjmed/hcs204

35. Zacharski LR, DePalma RG, Shamayeva G, Chow BK. The Statin–Iron Nexus: Anti-Inflammatory Intervention for Arterial Disease Prevention. Am J Public Health [Internet]. 2013 Apr [cited 2024 Oct 21];103(4):e105. Available from: https://pmc.ncbi.nlm.nih.gov/articles/PMC3673278/

36. Yu S, Chu Y, Li G, Ren L, Zhang Q, Wu L. Statin Use and the Risk of Cataracts: A Systematic Review and Meta-Analysis. J Am Heart Assoc Cardiovasc Cerebrovasc Dis [Internet]. 2017 Mar 20 [cited 2024 Oct 21];6(3):e004180. Available from: https://pmc.ncbi.nlm.nih.gov/articles/PMC5523994/

37. Bowden J, Madsen J, Goldman B, Iversen AT, Liang X, Vansteelandt S. Instrumental Variable methods to target Hypothetical Estimands with longitudinal repeated measures data: Application to the STEP 1 trial [Internet]. arXiv; 2024 [cited 2024 Dec 3]. Available from: http://arxiv.org/abs/2407.02902

38. 38. Bowden J, Pilling LC, Türkmen D, Kuo CL, Melzer D. The Triangulation WIthin a STudy (TWIST) framework for causal inference within pharmacogenetic research. Kutalik Z, editor. PLOS Genet [Internet]. 2021 Sep 8 [cited 2023 Jan 25];17(9):e1009783. Available from: https://dx.plos.org/10.1371/journal.pgen.1009783

39. Alver M, Kasela S, Haring L, Luitva LB, Fischer K, Möls M, et al. Genetic predisposition and antipsychotic treatment effect on metabolic syndrome in schizophrenia: a ten-year follow-up study using the Estonian Biobank. Lancet Reg Health - Eur [Internet]. 2024 Jun [cited 2024 Nov 20];41:100914. Available from: https://linkinghub.elsevier.com/retrieve/pii/S2666776224000814

40. Mayerhofer E, Malik R, Parodi L, Burgess S, Harloff A, Dichgans M, et al. Genetically predicted on- statin LDL response is associated with higher intracerebral haemorrhage risk. Brain J Neurol. 2022 Aug 27;145(8):2677–86.

41. Türkmen D, Masoli JAH, Kuo C, Bowden J, Melzer D, Pilling LC. Statin treatment effectiveness and the SLCO1B1 *5 reduced function genotype: Long-term outcomes in women and men. Br J Clin Pharmacol [Internet]. 2022 Jul [cited 2022 Dec 1];88(7):3230–40. Available from: https://onlinelibrary.wiley.com/doi/10.1111/bcp.15245

